# Ongoing natural selection drives the evolution of SARS-CoV-2 genomes

**DOI:** 10.1101/2020.09.07.20189860

**Authors:** Qi Liu, Shilei Zhao, Yali Hou, Sicheng Ye, Tong Sha, Yankai Su, Wenming Zhao, Yiming Bao, Yongbiao Xue, Hua Chen

## Abstract

SARS-CoV-2 is a new RNA virus affecting humans and spreads extensively through world populations since its first outbreak in late December, 2019. Whether the transmissibility and pathogenicity of SARS-CoV-2 is actively evolving, and driven by adaptation to the new host and environments is still unknown. Understanding the evolutionary mechanism underlying epidemiological and pathological characteristics of COVID-19 is essential for predicting the epidemic trend, and providing guidance for disease control and treatments. Interrogating 22,078 SARS-CoV-2 genome sequences of 84 countries, we demonstrate with convincing evidence that (i) SARS-CoV-2 genomes are overall conserved under purifying selection. (ii) Ongoing positive selection is actively driving the evolution of specific genes. Notably, genes related to coronavirus infection and host immune system defense are under adaptive evolution while genes related to viral RNA replication, transcription and translation are under purifying selection. A spatial and temporal landscape of 54 critical mutants is constructed based on their divergence among viral haplotype clusters, of which multiple mutants potentially conferring viral transmissibility, infectivity and virulence of SARS-CoV-2 are highlighted.

## Introduction

A newly emerged betacoronavirus, severe acute respiratory syndrome coronavirus 2 (SARS-CoV-2), causes a worldwide pandemic of the coronavirus disease 2019 (COVID-19), presenting a devastating threat to human public health attributed to its high infectivity and fatality (1–4). As of 18 July 2020, COVID-19 has resulted in 14,060,402 worldwide confirmed infections and 601,820 deaths across 188 countries and regions (https://coronavirus.jhu.edu/map.html).

RNA viruses usually have high mutation rates and tend to evolve rapidly (5). For a new RNA virus affecting humans, such as SARS-CoV-2, a recent host shift likely decreases its fitness and impels the virus to adapt to the new host environments and public health interventions (6, 7). Natural selection may act on the transmissibility and virulence of SARSCoV-2 through adaptive dynamics of specific genomic mutations, which has been observed in Ebola, Zika and other viruses (6, 7). SARS-CoV-2 has circulated globally since its first outbreak in late December, 2019, and accumulated plenty of genetic polymorphisms within the short period. There are currently more than 90,000 complete SARS-CoV-2 genome sequences publicly available, of which 16,183 nucleotide substitutions have been identified (data retrieved from https://bigd.big.ac.cn/ncov/), compiling the largest population genomic data of non-humans so far. These SARS-CoV-2 lineages have exhibited considerable variation of transmission and clinical characteristics in different countries or regions. For example, the basic reproduction number (R_0_) ranges between 2.2 and 5.9 (8, 9), and the mortality rate from 0.8% to 14.5% (10).

It is critical for epidemic trend prediciton and disease control to understand whether natural selection is actively driving the adaptive evolution of transmissibility and virulence of SARS-CoV-2 during the pandemic. If it is under selection, further research is warranted to identify the functional mutants contributing to the evolving epidemiological and pathogenic characteristics. Multiple studies have been carried out to explore the problem. Tang et al (2020) noticed that at the early stage of the COVID-19 outbreak, two major virus lineages (the S and L lineages) showed dramatic allele frequency difference before and after the lockdown of Wuhan city, and proposed that it may be due to natural selection favoring the spread of less severe lineages (11). However the argument is not conclusive for that allele frequency change is more likely caused by other confounding factors, e.g., stochastic fluctuation, founder effect or sampling bias (7, 12). Another study found the D614G replacement on one receptor binding domain of the Spike protein is predominant in multiple regions or countries and associated with higher upper respiratory tract viral loads, indicating a possible fitness advantage (10). Nevertheless, all these studies are based on allele frequency increase or differentiation that is not necessarily due to natural selection. It is only useful for screening for the putative mutant loci instead of serving as a persuasive proof of the hypothesis. Despite the individual functional mutants identified in aforementioned studies, “there is a lack of compelling evidence” of mutations that “impact the progression, severity or transmission of COVID-19 in an adaptive manner” (7). Indeed, the evolutionary driving forces underlying the epidemiological dynamics and pathogenic changes of SARS-CoV-2 remain elusive and uncertain. Furthermore, a genome-wide survey of functional mutations and their implication of the epidemiological perspective were not fully accomplished either.

Herein, we analyze SARS-CoV-2 genome sequences currently collected from different geographies and time points of the outbreaks. We utilize patterns of genomic polymorphism based on the ratio of nonsynonymous mutations (N) vs synonymous mutations (S) in different groups, which are exclusively caused by natural selection and robust to clustering infections, founder effects and sampling bias commonly existing in viral genomic data. The analysis validates the hypothesis that ongoing natural selection is actively acting on the SARS-CoV-2 genomes, shaping the epidemic dynamics of COVID-19. We then partition the viral lineages of world samples into clusters according to their genomic similarity as a result of global transmission and clustering outbreaks. A spatial and temporal landscape of mutants is constructed based on the clusters, and critical mutants potentially conferring pathogenic and clinical characteristics of SARS-CoV-2 are highlighted. Our results provide a reference of genomic mutations for epidemic prediction, surveillance and clinical treatments of COVID-19.

## Results

22,078 SARS-CoV-2 genome sequences publicly available from 84 countries as of June 2, 2020, across Asia, Europe, America, Africa, and Oceania were downloaded and aligned using MUSCLE (13). The 5’- and 3’- untranslated regions (UTRs) and regions with a high missing rate were trimmed out, with a final length of 29,410 nucleotides remained. A total number of 8,999 mutations were identified.

Since the virus populations are undergoing multiple clustering infections and founder effects, it is hard to use allele frequencies to draw reliable conclusions on virus evolution (12). On the premise of neutral evolution, the relative numbers (ratio) of non-synonymous mutants (N) vs synonymous mutants (S) should be a constant unaffected by changes of population sizes, sine both S and N sites should demonstrate identical population behavior under the common demographic history and neutrality. Accordingly in the following sections, our analysis is mainly based on the analysis of N and S numbers for different sets of mutations.

### Purifying selection dominates the viral genomic evolution

We first calculate the ratio of N/S using only mutation sites in very low frequencies. 4,573 non-synonymous mutations and 2,202 synonymous mutations with the minor frequencies (f) < 0.01% are identified. The N/S ratio is calculated to be 2.0768. From population genetics theory we know that when in very low frequencies, the population behavior of deleterious or beneficial mutants is essentially the same as that of selectively neutral mutants (14). Therefore, the N/S ratio 2.0768 represents the relative abundance of N and S mutations when the genome evolves under neutrality, and is determined only by sequence composition and nucleotide substitution rates of viral genomes. In the following analysis, we use the N/S ratio of mutations with f < 0.01% as the reference under neutral conditions.

We then check the relative occurrences of N and S mutations with higher frequencies. As we can see in Table 1, the ratio of N/S for mutations with the frequency f>0.001 decreases to1.52 and is significantly lower than N/S for f< 0.01% (P = 0.0034, Chi-squared test), indicating a negative selection against non-synonymous mutations with f>0.001 during the viral evolution. We further carry out the same analysis for subsets of mutations with different allele frequency ranges (e.g., (0.0001, 0.001], (0.001, 0.01], (0.01, 0.05]). The pattern of reduced N/S ratio is consistently observed, and the N/S ratio value keeps decreasing with the increase of f. The N/S ratio is 1.557 for SNPs with 0.0001 < f < = 0.001, is 1.50 for 0.001 <f < = 0.01, and 1.46 for 0.01 <f < = 0.05 (see Table 1). It is consistent with the fact that mutations with higher allele frequencies are normally older than those with lower allele frequencies and could be under natural selection for a longer duration showing more prominent effect.

**Table 1.**
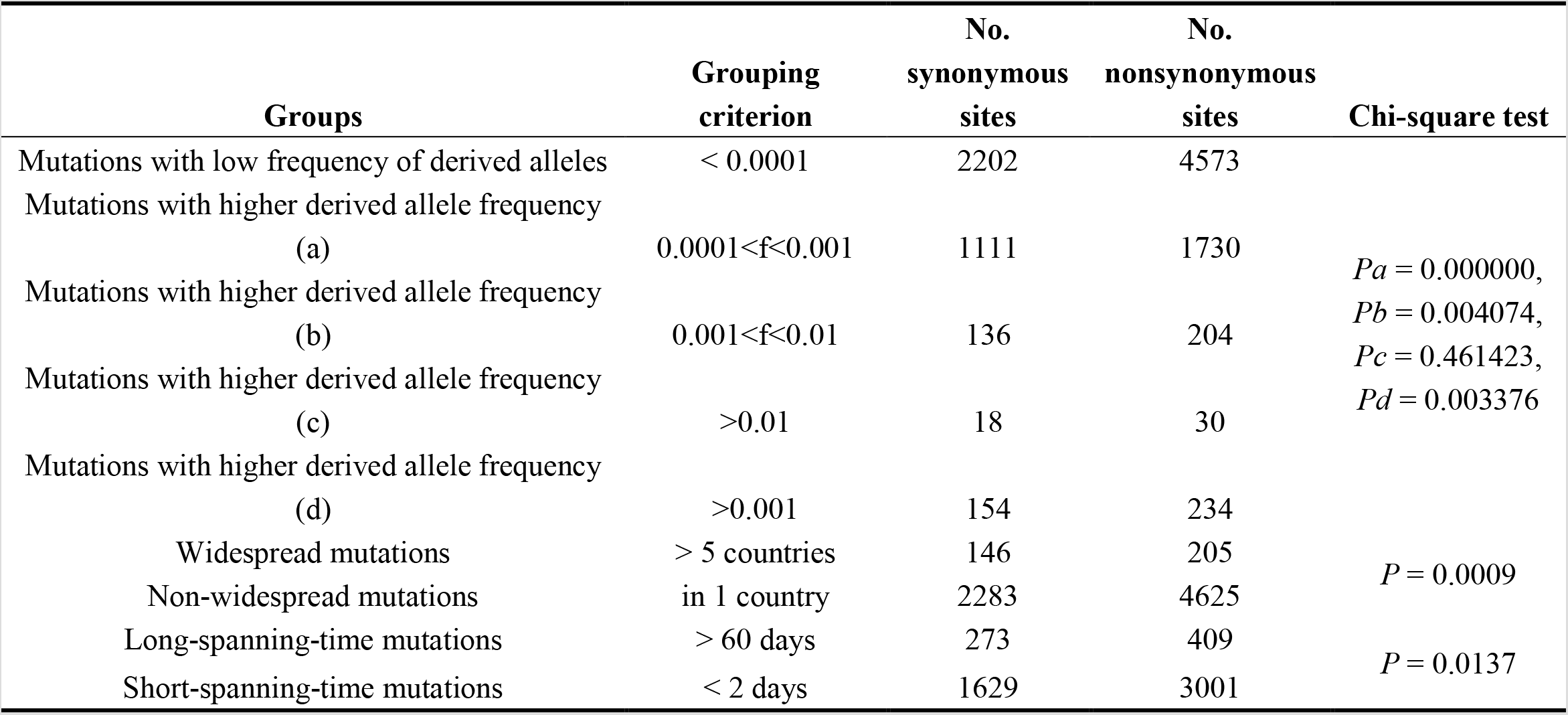
Chi-squared tests to compare the N/S ratios between different mutation groups.

We further compare numbers of synonymous and nonsynonymous mutations for widespread (defined as mutants observed in samples of more than 5 countries) and non-widespread (defined as mutants observed only in 1 country) variations. The widespread mutations tend to exist in the populations for a longer time than non-widespread ones. The N/S ratio of mutants predisposed to spread more countries is significantly lower *(P* = 0.0009, Chi-squared test; Table 1), suggesting that purifying selection acting on widespread mutations which exist for longer time and resulting a lower N/S ratio. We also group mutations according to the spanning of their sample-collection times, which is calculated as the durations between the earliest and most recent sample-collection times of viral genomes carrying that mutations. The N/S ratio of short-spanning-time mutations (< 2 days) is 1.842 (3001/1629), while the N/S ratio of long-spanning-time mutations (>60 days) is 1.498 (409/273). The lower N/S ratio indicates more selective constrains on these long-spanning-time mutations, again, confirming the effect of purifying selection (P = 0.0137, Chi-squared test; Table 1).

In summary, the aforementioned three analyses reveal that despite of spreading for more than eight months and the accumulation of sufficient mutations, the SAR-CoV-2 genomes are overall under purifying selection and relatively conserved. The conclusion also accords with the low genome-wide mutation rate inferred recently (0.000869 per site per year (12)).

### Positive selection drives the adaptive evolution of genes conferring pathogenicity and infectivity

Even though SARS-CoV-2 are under genome-wide negative selection, a small fraction of the viral genome may have undergone positive selection, resulting in pathogenic and clinical variety across geographical regions, of which the genetic polymorphism pattern may be diluent in the genome-wide N/S ratios. To detect positive selection on a specific gene, we perform Chi-squared tests by comparing the ratio of nonsynonymous versus synonymous mutations at individual gene level with the ratio from genome-wide mutants with the derived allele frequency f < 0.0001, which are assumed to be identical to under neutral evolution.

Chi-squared tests indicate that three genes including the open reading frame 3 *(ORF3a)*, *ORF8*, and *ORF10* show significantly a higher proportion of nonsynonymous mutations *(P* < 0.05; Fig. 1; Table 2), presenting evidence of positive selection. Another gene encoding the nucleocapsid *(N)* protein also shows an elevated N/S ratio (2.422) with the P-value close to 0.05 (*P* = 0.065). Interestingly, except for the *ORF10* gene that has been under very limited studies, the rest three positively-selected genes encode viral accessory proteins that play critical roles in virus-host interactions and host immune response modulation, contributing to different pathogenic outcomes (15, 16). *ORF3a* has been demonstrated to induce cellular apoptosis that is recognized to antagonize host antiviral immune defense system and facilitate viral pathogenicity (16); *ORF8* was reported to disrupt antigen presentation and mediate immune evasion that warrants further investigation (17, 18); *N* protein, directly binding to viral RNA, is essential for viral encapsidation, and was also reported participating in immune-evasion mechanism and promoting viral infections (19–22). These findings highlight that during COVID-19 global pandemic, positive selection is very likely an essential driving force and is acting on the interplay between coronavirus infection and host immune system defense.

**Figure 1.**
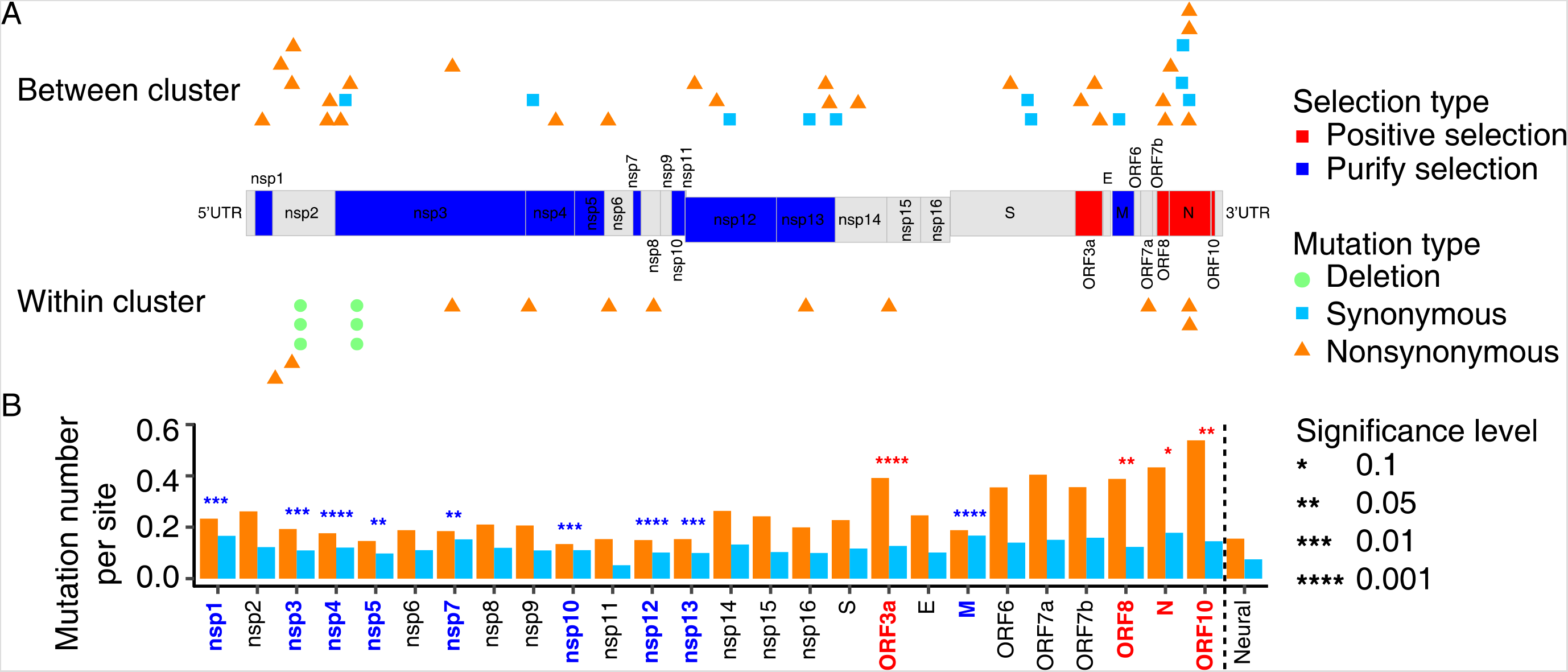
Evidence of natural selection acting on the SARS-CoV-2 genome. (A) Genes significantly under positive selection are in red, and those under negative selection in blue. The putatively selected mutations of each gene are presented (upper panel: between- cluster mutants; lower panel: within-cluster mutants). Deletions are denoted with circles, synonymous variations are with squares, and nonsynonymous variations with triangles. (B) The numbers of synonymous (light blue) and nonsynonymous (orange) substitutions per site for each gene. Significant levels of the excess of nonsynonymous versus synonymous mutations for each gene were achieved by using a Chi-squared test and comparing to those from rare mutations with the allele frequency < 0.0001.

**Table 2.** The N/S ratio and function annotation of each gene, and the corresponding p values of Chi-squared test indicating whether they are under natural selection.

| Gene name | Non-synonymous site number | Synonymous site number | N/S ratio | p value | Gene function |
| --- | --- | --- | --- | --- | --- |
| **Genes with a N/S ratio greater than neutrality (N/S > 2.0767)<sup>a</sup>** |  |  |  |  |  |
| *ORF3a* | 324 | 105 | 3.0857 | 0.000549 | Inducing apoptosis |
| *ORF8* | 142 | 45 | 3.1556 | 0.014909 | disrupting antigen presentation and mediates immune evasion |
| *ORF10* | 63 | 17 | 3.7059 | 0.032489 | Unknown |
| *N-protein* | 545 | 225 | 2.4222 | 0.064752 | Contribute to viral genome assembly and serve as viral suppresor of RNAi to antagonize the host immune defense system |
| ORF7a | 148 | 55 | 2.691 | 0.104571 | Putative type I transmembrane protein |
| NSP15 | 252 | 107 | 2.3551 | 0.287196 | Endoribonuclease and potential degrading viral dsRNA to prevent host recognition |
| S-protein | 870 | 447 | 1.9463 | 0.308559 | Glycoprotein mediating attachment of the virus to the host cell |
| ORF6 | 66 | 26 | 2.5385 | 0.388119 | Potentail interacting with NSP8 to promoting RNA polymerase activity and contribute to viral pathogenesis |
| E-protein | 56 | 23 | 2.4348 | 0.52257 | Small integral membrane protein involving in viral replication cycle, viral assembly, virion release and viral pathogenesis |
| NSP14 | 417 | 210 | 1.9857 | 0.612499 | 3' to 5' endonuclease and N7-methyltransferase |
| NSP9 | 70 | 37 | 1.8919 | 0.649014 | RNA binding protein |
| NSP11 | 6 | 2 | 3 | 0.650707 | Unknown |
| NSP2 | 500 | 235 | 2.1277 | 0.771068 | Binding to prohibitin 1 and prohibitin 2 (PHB1 and PHB2) and involving in disrupting the host cell environment |
| NSP16 | 178 | 89 | 2 | 0.776064 | 2'-O-Ribose-Methyltransferase |
| ORF7b | 47 | 21 | 2.2381 | 0.77661 | Unknown |
| **Genes with a N/S ratio smaller than neutrality (N/S < 2.0767)<sup>a</sup>** |  |  |  |  |  |
| ***M-protein*** | **126** | **112** | **1.125** | **0.000003** | Contribute to the shape of the virus envelope and promotes completion of viral assembly |
| ***NSP12*** | **419** | **282** | **1.4858** | **0.000036** | RNA-dependent RNA polymerase involving in viral replication |
| ***NSP4*** | **265** | **181** | **1.4641** | **0.000439** | Transmembrane scaffold protein |
| ***NSP13*** | **276** | **179** | **1.5419** | **0.002658** | RNA helicase, 5' triphosphatase |
| ***NSP3*** | **1126** | **639** | **1.7621** | **0.003208** | Promoting cytokine expression and cleavage of viral polyprotein |
| ***NSP1*** | **126** | **90** | **1.4** | **0.004733** | Leader protein, Inhibition of IFN signaling and blocking of host innate immune response by promotion of cellular degradation and blocks translation of host's RNA |
| ***NSP10*** | **56** | **46** | **1.2174** | **0.007106** | Stimulation of exoribonuclease and 2'-O-methyltransferase activity |
| ***NSP7*** | **46** | **38** | **1.2105** | **0.013368** | Processivity clamp for RNA polymerase by arms hexadecameric complex |
| ***NSP5*** | **134** | **90** | **1.4889** | **0.016007** | Promoting cytokine expression and cleavage of viral polyprotein |
| NSP6 | 164 | 96 | 1.7083 | 0.13577 | Putative transmembrane protein and potential limiting autophagosome/lysosome expansion |
| NSP8 | 125 | 71 | 1.7606 | 0.273062 | Processivity clamp for RNA polymerase by arms hexadecameric complex |

(<sup>a</sup>: Mutations with the derived allele frequency < 0.0001 were used as neutral control. The number of nonsynonymous mutations: 4573; the number of synonymous mutations: 2202; N/S ratio: 2.0767.)

In contrast, the majority of the viral genes demonstrates a N/S ratio < 2.0 (Fig. 1, Table 2), being consistent with last section that purifying selection is dominant. Nine genes show statistically significantly reduced proportion of nonsynonymous substitutions *(P* < 0.05; Table 2). Eight of the negatively selected genes *(NSP12, NSP4, NSP13, NSP3, NSP1, NSP10, NSP7* and *NSP5)* encode non-structure proteins of SARS-CoV-2, including the main protease, papainlike protease, RNA helicase, RNA-dependent RNA polymerase, and so on, which are essential in viral RNA replication, transcription, and translation (23). In addition, the most significant gene under negative selection encodes the SARS-CoV-2 membrane *(M)* protein that plays a central role in viral assembly and viral particle formation (23). The results indicate that proteins conferring fundamental molecular functions, such as viral replication, translation etc., are under significant purifying selection.

### Clustering pattern of viral lineages

As we have demonstrated, although the viral genomes are under purifying selection, positive selection has been driving the evolution of genes related to coronavirus infection and host immune system defense. A further target is to identify the putative functional mutations under selection. Some recent studies have provided a list of candidate mutations, most of which were identified using allele frequency changes in the overall global samples. As we know, the viral populations have been evolving and spreading in heterogeneous rates, demonstrating a clustering pattern. Investigating the allele frequencies in the pooled sample from multiple populations has two limitations: first, it has limited power to identify mutants which arose in a local population recently while are in a low frequency in the global population; second, it provides little information on the spatial and temporal origin (when and where) of these functional mutations. To track the evolutionary dynamics of genomic variants in a fine scale, we partition the sample of 22,078 genomes into distinct clusters according to their sequence similarity and evolutionary relationship, and identify 14 worldwide predominant clusters of SARS-CoV-2, denominated as C01, C02, …, and C14, respectively (see Fig. S1 and the Methods section for details of the partition approach). The 14 clusters and their genealogical relationship on a haplotype network are presented in Fig. 2A, and spatially arranged with their x-axis given by the earliest sampling dates of viral samples from each cluster (Fig. 2A).

**Figure 2.**
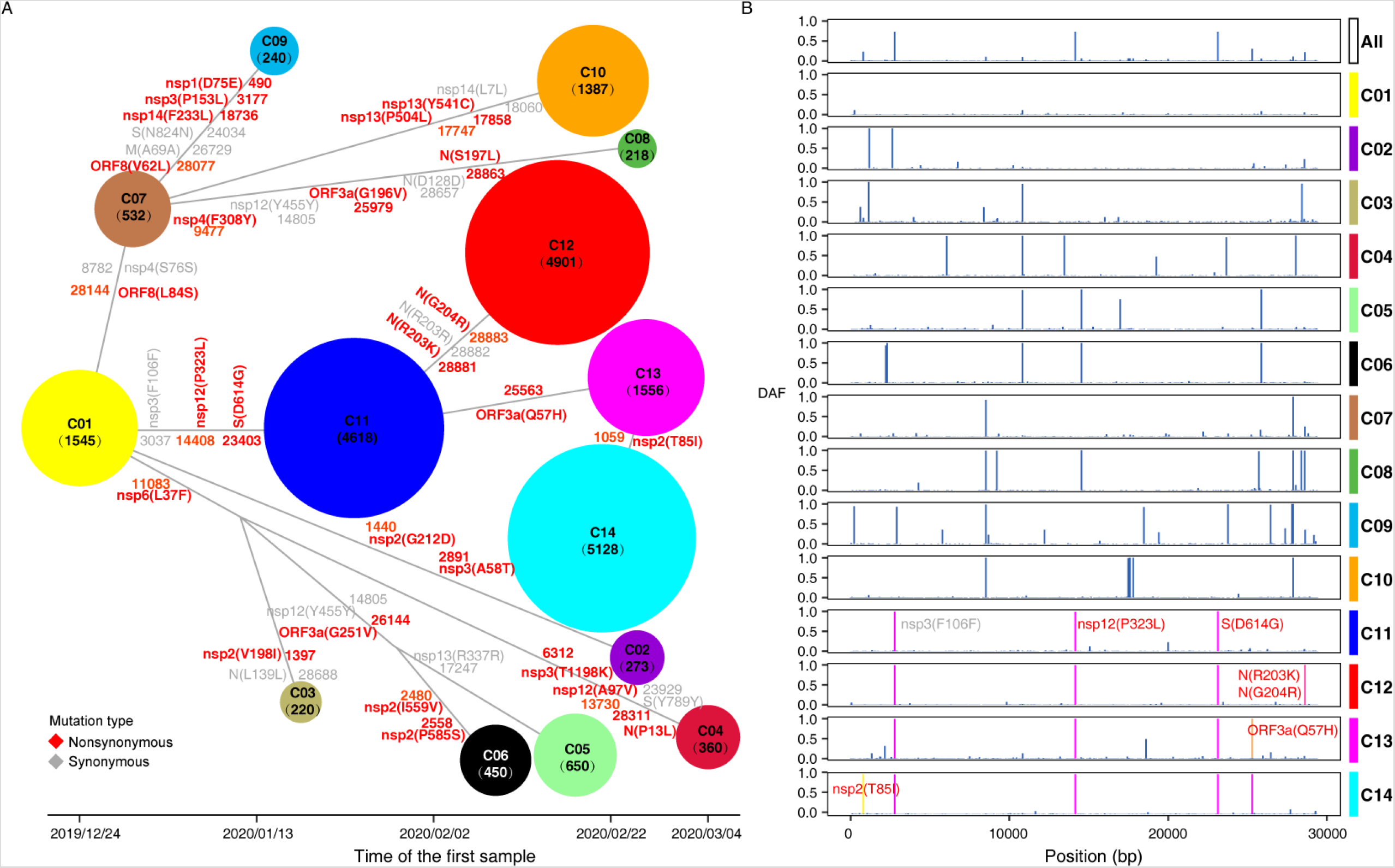
Fourteen major worldwide haplotype clusters of SARS-CoV-2 presented according to their genealogical relationship and the earliest sampling dates of the clusters along the x-axis. (A) The x-axis represents the sampling date of the first collected sample belonging to each cluster. The nodes represent different haplotype clusters, with the node sizes proportional to the counts of the belonged sequences (ranging from 218 to 5,128 sequences). Mutations that are highly differentiated between clusters are marked on the branches (1 ∼ 6 mutations per branch), of which nonsynonymous and synonymous variations are in red and gray respectively. The positions of the mutations are from the genome reference of WH-Hu-1 (NC_045512.2). When the SNP has multiple alleles (> 2), the most frequent derived allele is selected and shown. (B) The spectrum of derived allele frequencies of mutations for each cluster. Mutations distinguishing the C11-C14 clusters are highlighted in red.

The 14 clusters correspond to infection clusters or major founder effects during the COVID-19 pandemic (12). C01 cluster is predominantly prevalent in East and Southeast Asian countries with the earliest sample collection date (December 24, 2019). C07 comprises another major haplotype type in East and Southeast Asia, as well as some African countries like Nigeria and Uganda. C03 primarily ravages in the countries mainly located in Western Asia. Viruses of C04 preponderate in Southeast Asia and India. C06 and C08 clusters are in Jamaica, Kazakhstan and Spain. C10 is in North America.

C11-C14 are the most abundant and widely distributed haplotype clusters in the sample. C11 broadly ravages 6 African, 26 European, 5 Latin American, and 2 Oceanian countries, as well as Canada, India and Russia. Similarly, C12 is primarily dominant in Europe, Latin America, Asia, and Africa, and C14 preponderates in Denmark, Israel, United States, and moderately occupies 11 European countries (such as Norway, Austria, France and Germany). C13 dominates in West Asian countries, and presents moderate frequencies in Turkey, Colombia, Chile and Israel etc. The remaining C02, C05, and C09 clusters represented small haplotype groups across broad countries.

### Tracking the spatial and temporal occurrence of putatively selected mutations along the pandemic dynamics

Nucleotide mutations that are predominant in a cluster and absent or in low frequencies in others are potentially of functional importance for virus pathogenicity and transmissibility, serving as targets of positive selection. Following this criterion, we identify 37 protein-coding variations differentiated among the 14 delineated haplotype clusters, including 26 nonsynonymous and 11 synonymous sites, among which 13 (12 nonsynonymous and 1 synonymous) are clade-defining loci with the allele frequency greater than 0.99 in one specific cluster (Table 3). We map the occurrence of these mutations to branches connecting the clusters on the haplotype network (see Fig. 2A). The numbers of inter-cluster mutants per branch vary from 1 to 6.

**Table 3.** The list of mutations with high among-cluster divergence that are potentially under positive selection.

| Cluster | Position | Gene | Substitution<br>change | Substitution<br>type | Mutation function | Cluster<br>classification<br>site |
| --- | --- | --- | --- | --- | --- | --- |
| C2 | 1440 | nsp2 | G212D | N | Located at homologous domain (PDB 3ld1) important for viral pathogenicity | 1 |
| C2 | 2891 | nsp3 | A58T | N | Located at N-terminal ubiquitin-like domain that plays an important role in viral replication |  |
| C3 | 1397 | nsp2 | V198I | N | Located at homologous domain (PDB 3ld1) important for viral pathogenicity |  |
| C3 | 28688 | N | L139L | S |  |  |
| C4 | 6312 | nsp3 | T1198K | N | Involving acquisition of a charged group |  |
| C4 | 13730 | nsp12 | A97V | N | Located at NiRNA domain involving the interaction with beta-hairpin of nsp12 protein | 1 |
| C4 | 23929 | S | Y789Y | S |  |  |
| C4 | 28311 | N | P13L | N | Might involving in viral replication |  |
| C5 | 17247 | nsp13 | R337R | S |  |  |
| C6 | 2480 | nsp2 | I559V | N | Might related with viral pathogenicity | 1 |
| C6 | 2558 | nsp2 | P585S | N | Might related with viral pathogenicity |  |
| C8 | 9477 | nsp4 | F308Y | N | Might related with viral replication | 1 |
| C8 | 25979 | ORF3a | G196V | N | Might involving in viral spread and pathogenesis |  |
| C8 | 28657 | N | D128D | S |  |  |
| C8 | 28863 | N | S197L | N | Located at the linker between NTD and CTD of N protein and might involving in viral replication |  |
| C9 | 490 | nsp1 | D75E | N | Might related with inhibition of host gene expression |  |
| C9 | 3177 | nsp3 | P153L | N | Might related with the proteolytic cleavage |  |
| C9 | 18736 | nsp14 | F233L | N | Located at N-terminal exonuclease domain of nsp14 |  |
| C9 | 24034 | S | N824N | S |  |  |
| C9 | 26729 | M | A69A | S |  |  |
| C9 | 28077 | ORF8 | V62L | N | Negatively modulating ORF8 interactions with C3b and might impacting host immune responses | 1 |
| C10 | 17747 | nsp13 | P504L | N | Increasing affinity of helicase RNA interaction | 1 |
| C10 | 17858 | nsp13 | Y541C | N | A destabilizing mutation increasing the molecular flexibility and decreasing affinity of helicase binding with RNA |  |
| C10 | 18060 | nsp14 | L7L | S |  |  |
| C3,C4,C5,C6 | 11083 | nsp6 | L37F | N | Affecting the protein-protein interactions | 1 |
| C5,C6 | 26144 | ORF3a | G251V | N | Possible involving the loss of a B-cell like epitope in ORF3a and viral spread |  |
| C5,C6,C8 | 14805 | nsp12 | Y455Y | S |  | 1 |
| C7,C8,C9,C10 | 8782 | nsp4 | S76S | S |  |  |
| C7,C8,C9,C10 | 28144 | ORF8 | L84S | N | Negatively modulating ORF8 interactions with C3b and might impacting host immune responses | 1 |
| C12 | 28881 | N | R203K | N | Predicted to reduce the binding of an overlying putative HLA-C*07-restricted epitope |  |
| C12 | 28882 | N | R203R | S | Predicted to reduce the binding of an overlying putative HLA-C*07-restricted epitope |  |
| C12 | 28883 | N | G204R | N |  | 1 |
| C14 | 1059 | nsp2 | T85I | N | Might involving in disrupting the host cell environment | 1 |
| C13,C14 | 25563 | ORF3a | Q57H | N | Might involving in viral spread and pathogenesis | 1 |
| C11,C12,C13,C14 | 3037 | nsp3 | F106F | S |  |  |
| C11,C12,C13,C14 | 14408 | nsp12 | P323L | N | Possible involving RNA-dependent-RNA polymerase fidelity | 1 |
| C11,C12,C13,C14 | 23403 | S | D614G | N | Associated with higher infectious titers of spike-pseudotyped virus,<br>and potentially higher viral loads in COVID-19 patients but not<br>with disease severity. |  |

C01, the cluster preponderates in East and Southeast Asia is likely the earliest cluster if using sample-collection dates as a criterion (also with the inferred TMRCA, results not shown). A nonsynonymous mutation L84S in the *ORF8* protein, together with a tightly linked synonymous variant (S76S) in the *NSP4* protein, emerged on the branch leading to the other primary cluster C07 in Asia (Fig. 2A). The L84S replacement together with S76S were used in former studies to define two major haplotype groups in the early epidemic stage: the L and S lineages. The proportion of L lineage in samples collected before and after Wuhan lockdown showed distinct differentiation (99% vs 70%), and it was hypothesized that the frequency change of L lineage may due to different pressures of negative selection from containment measures (11). It is disputable for that the allele frequency change can be caused by sampling bias and clustering infection as well (12). In the map of haplotype network, L84S is pinpointed to the branch connecting C01 and C07 (Fig. 2), with a very low frequency of 0.1294% in C01 and increased to 100% in C07, demonstrating an obvious cluster expanding pattern in the fine scale. Furthermore, we identified the signature of positive selection acting on *ORF8* as mentioned previously. Based on the two facts, this mutation is possibly a candidate site under positive selection for further evaluation. According to COVID-3D database, the L84S amino acid alteration was predicted to eliminate 4 hydrophobic bonds and lead to destabilization of *ORF8* protein. Another study with computational protein modeling proposed that L84S can mitigate the binding of *ORF8* to human complement *C3b*, which is negatively regulated by the C-terminus serine-protease catalytic domain of the human complement factor 1 *(F1)*, and activates the host complement system (24). Therefore, the L84S mutation possibly impacts the normal function of *ORF8*, and plays an important role in the host immune responses and infection outcome.

A recurrent nonsynonymous mutation D614G in the epitope region (the receptor-binding domain) of Spike glycoprotein protein *(S)* occurred in the common ancestor of the C11-C14 clusters (Fig. 2A). C11-C14 clusters were widely preponderated across Europe, America, Asia, and Africa. Spike glycoprotein is essential for the host cell infection of SARS-CoV-2. D614G was reported to be relevant to a high viral load in COVID-19 patients and higher infectious titers of spike-pseudotyped viruses, indicating a possible fitness advantage (10). Another non-synonymous mutation P323L in *NSP12* (RNA-dependent RNA polymerase, RdRP) also occurred in the common ancestor of C11-C14, coupling the evolution with D614G. This mutation might regulate the activity of RdRp, and is related to viral replication and fidelity (25).

Derived from C11, two non-synonymous mutations, R203K and G204R on the phosphoprotein domain of *N* protein arose to 100% in C12. Both clusters are widely distributed in European countries, besides, C11 is predominant (>50%) in Africa (Congo, South Africa, and Kenya), while C12 is in Vietnam, Latin America (Brazil and Argentina), Bangladesh, and Russia. According to structural prediction provided by the COVID-3D database, R203K and G204R both destabilize the *N* protein, and the predicted actual free energy value (ΔΔG) using the mutation Cutoff Scanning Matrix (ΔΔG^stability^ mCSM, henceforth) are −1.71 and –1.07 kcal.mol^−1^ respectively, resulting the alteration of their molecular interactions with other amino acids, such as carbonyl, polar bonds, and hydrogen bonds (Fig. 3A-D). Mutations on *N* protein that was under positive selection may be functional relevant to viral replication, assembly and participating in immune-evasion and viral infections (19–22). We further infer the mortality rates of viruses belonging to the two clusters (see Methods). The mortality rates of C11 and C12 are dramatically different (16.03% and 3.98%), and may be explained by the functional alternation of *N* protein caused by the two nonsynonymous mutations.

**Figure 3.**
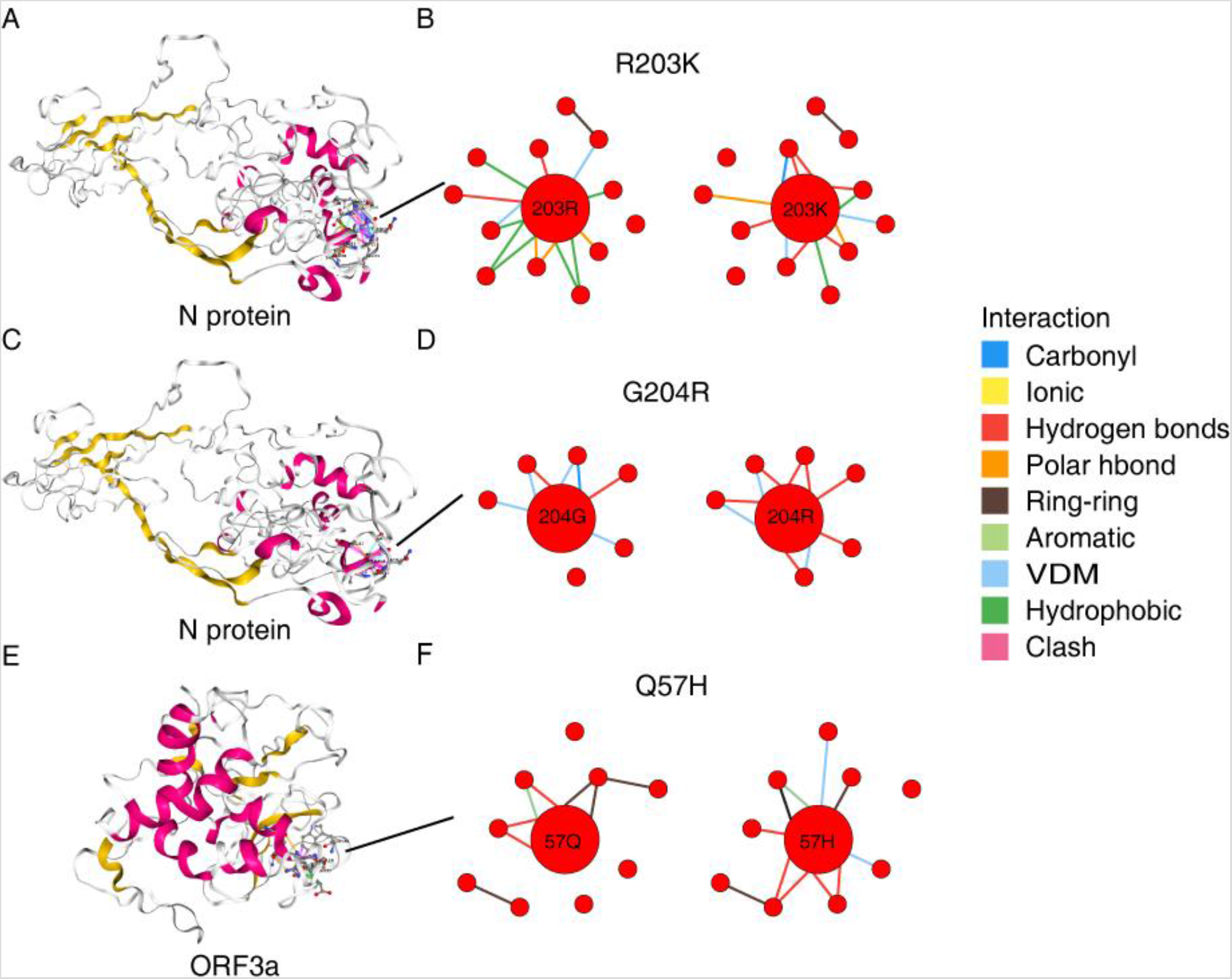
The impact of mutations on protein structures. (A), (C) The predicted 3D structure for nucleocapsid *(N)* protein. (B) alteration of molecular interactions by introducing the R203K mutation. (D) The alteration of molecular interactions by introducing the G204R mutation. (E) The predicted 3D structure for *ORF3a* protein. (F) The alteration of molecular interactions by introducing the Q57H mutation.

Diverged from C11, the Q57H mutation in *ORF3a*, another positively selected gene, arose and fixed in the C13 cluster that dominates in Saudi Arabia and broadly distributed in Europe (Fig.2). This variant was predicted to exert structural destabilization (ΔΔG^stability^ mCSM = –1.55 kcal.mol^−1^) (Fig. 3E-F) and deleterious effects on protein function (Issa et al., 2020). Another mutation T85I in *NSP2* further diverges C14 from C13 that preponderates (>50%) in USA and European countries and with a lower mortality rate (2.83%). This mutation also fall within the region of *NSP2* homologous to the protein (PDB 3ld1), and may be functional in viral pathogenicity (28).

A full list of the 37 mutations and their functional significance can be found in Table 3.

### Mutations with rapid increase of frequency within clusters as potential targets

Other than the mutations demonstrating nearly-fixed divergence among clusters, mutations presenting prominent frequency increasing trend over sampling times within a cluster are potential sites with evolutionary or functional significance in COVID-19 epidemic. We profile those within-cluster mutations according to their frequency dynamics during a period of 5 sequential sampling times. 17 mutations demonstrating a trend of frequency increase over sampling times were identified (Table 4).

**Table 4.** The list of mutations with rapid increase of frequency within clusters as potential targets of positive selection.

| Position | Gene | Substitution<br>change | Substitution<br>type | Mutation function |
| --- | --- | --- | --- | --- |
| 1605 | nsp2 | N267Del | Deletion | Located at homologous domain (PDB 3ld1) important for viral pathogenicity |
| 1606 | nsp2 | N267Del | Deletion | Located at homologous domain (PDB 3ld1) important for viral pathogenicity |
| 1607 | nsp2 | D268Del | Deletion | Located at homologous domain (PDB 3ld1) important for viral pathogenicity |
| 884 | nsp2 | R27C | N | Located at homologous domain (PDB 3ld1) important for viral pathogenicity |
| 3333 | nsp3 | I205Del | Deletion | Located near the domain (ADRP) important for viral replication |
| 3334 | nsp3 | I205Del | Deletion | Located near the domain (ADRP) important for viral replication |
| 3335 | nsp3 | E206Del | Deletion | Located near the domain (ADRP) important for viral replication |
| 8653 | nsp4 | M33I | N | Might related with resistance to nucleoside analog 2' ,3' -dideoxy-3' thiacytidine (3TC) |
| 17141 | nsp13 | A302D | N | Might related with helicase activity and viral translation |
| 6310 | nsp3 | S1197R | N | Located at homologous nucleic acid-binding domain (PDB 2K87) |
| 28878 | N | S202N | N | Located at the linker between NTD and CTD of N protein and might involving in viral replication |
| 12478 | nsp8 | M129I | N | Might related with RNA-dependent RNA polymerase activity |
| 19684 | nsp15 | V22L | N | Might related with preventing host recognition |
| 27635 | ORF7a | S81L | N | Might related with ORF7a-dependent induction of apoptosis |
| 28896 | N | A208G | N | Located at the linker between NTD and CTD of N protein and might involving in viral replication |
| 1401 | nsp2 | G199E | N | Located at homologous domain (PDB 3ld1) important for viral pathogenicity |
| 11109 | nsp6 | A46V | N | Located in second transmembrane helix of nsp6 |

Within the C1 cluster, 3 consecutive nucleotide deletions causing a frameshift and two amino acid changes in *NSP2* (from 267N & 268D to N) rise in frequency especially in United Kingdom during the period of 5 sequential sampling times (Table 4). This mutation appears in Australia in March 14, 2020 and its frequency arises subsequently. The variations locate in the region homologous to the endosome‐associated protein in avian infectious bronchitis virus that plays a key role in the viral pathogenicity (28).

Three variations in complete linkage disequilibria uniquely occurred within C3, especially in Kazakhstan after April, 2020 (Table 4). The variations include 2 linked nonsynonymous mutations *(NSP2* R27C and *NSP4* M33I), and a 3-nucleotide deletion in *NSP3* (3333T, 3334T, and 3335G) that results in a replacement of 205I & 206E with Lysine, as well as A302D substitution in the helicase domain of *NSP13* that potently suppresses primary interferon production and signaling thereby immune suppression (29).

A nonsynonymous mutations G199E of *NSP2* raises the frequency gradually from 0 to 0.5 within the C10 cluster across sampling dates (Table 4). The pattern occurred especially in the United States. G199E locates in the homologous region (PDB 3ld1) of *NSP2*, and may be functionally important in viral pathogenicity according sequence homologous analysis (28).

## Discussion

It is well known that viruses mutate and adapt rapidly to environments and hosts (30, 31). SARS-CoV-2 is a new RNA virus affecting humans and spreads extensively through world populations since its first outbreak in late December, 2019. The virus lineages have demonstrated spatial heterogeneity in transmissibility and infectivity. Whether the epidemical dynamics and pathogenicity of SARS-CoV-2 is actively driven by natural selection is still unknown. Understanding the underlying evolutionary mechanism is essential for elucidating the viral pathogenicity, predicting the epidemic trend, and providing guidance for disease control and treatments.

The dynamics of virus populations demonstrates a series of founder events caused by infection clustering or bursts of epidemic in local regions. Besides, the genomic samples are usually collected from different times and locations disproportionally (sampling bias). Both significantly impact the allele frequencies and bring challenges in analyzing the virus genomic data with most population gendetic methods (12). Comparing the relative excess of synonymous and nonsynonymous substitutions is robust to population size changes, representing an efficient approach for evaluating the effects of natural selection on SARS-CoV-2. The approach we used in this study is similar to the known McDonald-Kreitman test (MK test) in molecular evolution, which compares the ratios of non-synonymous site number over synonymous site number of between between-species divergence to that of within-species polymorphisms, and uses the latter as an internal control under neutrality. However, the method we proposed here is for only comparing genetic polymorphisms within a species. The method is novel in using the N/S ratio of rare mutants with very low frequencies as the internal control under neutral evolution, which is valid according to population genetic theory (14). The method is only applicable for large-sample genomic sequencing data.

We demonstrate with multiple lines of evidence that SARS-CoV-2 genomes are overall constrained under purifying selection. Evidences of positive selection acting on specific genes are also observed. We have revealed that the genes encoding viral accessory proteins and *N* protein that participate in coronavirus infection and host immune response present signatures of positive selection; while *NSP* and *M* proteins implicated in viral RNA replication and transcription were shaped by purifying selection.

We further partition the viral genomic samples into 14 haplotype clusters according to their sequence similarity and genealogical relationship. Superimposing on the 14 worldwide transmission clusters, we provide a list of mutations as putative targets of natural selection. Whilst there is no concrete evidence supporting their functional significance during the outbreaks, mutations show between-cluster divergence or within-cluster frequency boost explain distinct pathogenicity and infectivity. Thus the list of mutations provides a basis for further functional study and clinical treatment.

## Material and methods

### SARS-CoV-2 genomes downloaded from public databases

SARS-CoV-2 genomic sequences were downloaded from two public databases, *i.e*., the 2019 Novel Coronavirus Resource (2019nCoVR, https://bigd.big.ac.cn/ncov/, (33)) and the Global Initiative on Sharing All Influenza Data (GISAID, https://www.gisaid.org/). The genome data set contains 22,243 genomic sequences from 84 countries with the sampling date ranging from December 23, 2019 to June 2, 2020. We filtered out 165 genomic sequences with missing on mutant loci that were used as features in haplotype clustering (see below), and 22,078 samples were retained for further analysis.

### Nucleotide mutation identification

All the sequences were aligned using MUSCLE (13) with default parameter settings. 264 bp of the 5’- untranslated region (UTR), 229 bp of the 3’-UTR region, and 1,646 bp of other regions with a missing rate > 99% in the alignment were trimmed out, finally retaining a total length of 29,410 nucleotides. Alleles of mutations homologous to the major allele in early virus strains (before March 1, 2020) were considered as the ancestral allele, otherwise as the derived allele.

### Partition haplotypes into clusters

The haplotype network of the viral samples demonstrates a multi-cluster pattern. The haplotype network is continuously divided into separate groups following steps of the decision tree shown in Figure S1. 14 SNPs are chosen as the features for classification. Each of the haplotypes are assigned to different clusters according to their alleles with the largest MAF (minor allele frequency) on the 14 SNP loci following the order of the features (Fig. S1). This process continued until no alleles with MAF > 0.1 or the subtree sample number < 500 (see classification rule in Fig. S1). 165 samples with missing data on the feature SNP locus are excluded from further analysis. After all samples are assigned to the clusters, we further implement an additional checking step by calculating the distance of all haplotypes to the MRCA sequences of the 14 clusters. We found that 185 samples (0.84% out of 22,078 sequences) are with a closer distance to a cluster other than their assigned clusters. These 185 samples were then re-assigned to the closest clusters.

### Prediction of the effects of mutations on protein function

We use COVID-3D (http://biosig.unimelb.edu.au/covid3d/), an online resource exploring the structural distribution of genetic variations of SARS-CoV-2, to predict the functional effects of mutations (24). It presents the change of energy of molecular structure and the difference in molecular interactions between wild-type and mutant-type alleles.

### Estimation of the lineage-specific mortality rates

We estimate the mortality rates of three prevalent clusters C11, C12 and C14 in European countries (C11, C12 and C14 account for 78.3 % of the samples from European countries). We filter data of different countries with two criteria: 1) sample size of the three clusters > 100; 2) number of cumulative infected > 10,000. The data of 12 remaining countries after filtering are used to estimate the mortality rate of the three clusters.

Let the mortality rates of C11, C12 and C14 lineages be *m*_1_, *m*_2_, *m*_3_ respectively. Denote the proportions of the three clusters in country *j* as 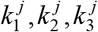. Denote the overall mortality rate of country *j* as *d_j_*. The mortality rate *d_j_* is thus a sum of *m*_1_, *m*_2_, *m*_3_ weighted by 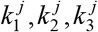. We have the overdetermined equations:

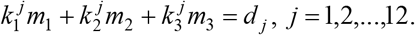

We solved the above sets of linear equations using the least square method implemented by the MATLAB built-in function *lsqlin*.

## Data Availability

All data are public data.

## Acknowledgements

We thank all the personal and institutions who have generated SARS-CoV-2 genome sequences, and GISAID (http://www.gisaid.org) and the National Genomics Data Center (https://bigd.big.ac.cn/ncov/) for sharing the SARS-CoV-2 data. The study was supported by the National Key R&D Program of China (Grant No. 2020YFC0847000, 2018YFC1406902, and 2018YFC0910402) and the Natural Science Foundation of China (Grant No. 31571370, 91731302, and 91631106).

---

**Figure S1.**
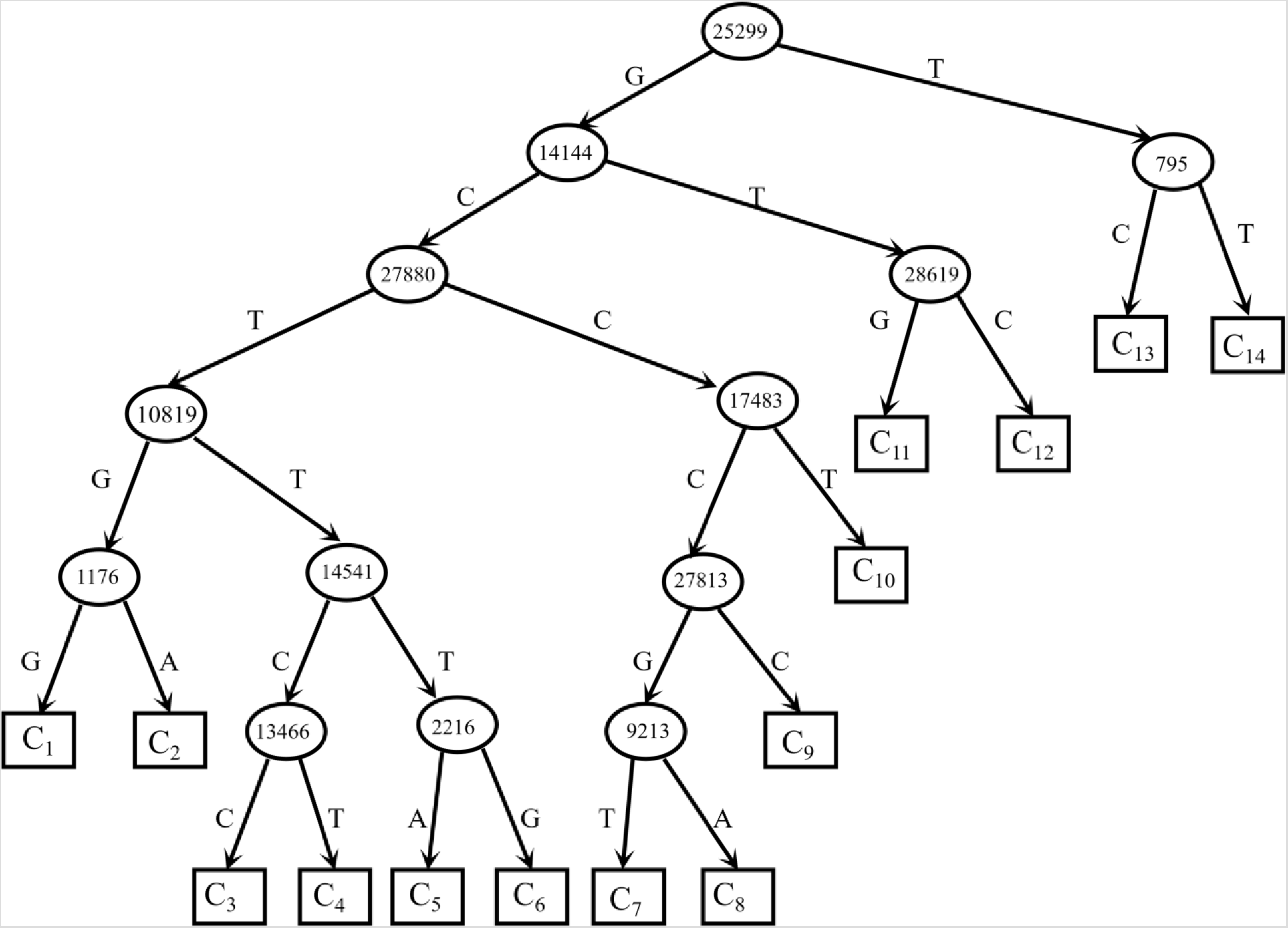
The classification tree and feature SNPs for assigning viral haplotypes to one of the 14 haplotype clusters.

## Notes

### Competing Interest Statement

The authors have declared no competing interest.

